# Impact of Tongan taxes targeting foods high in fat, salt and sugar: Interrupted time series analyses

**DOI:** 10.1101/2025.05.08.25327266

**Authors:** Andrea Teng, Viliami Puloka, Alice Hyun Min Kim, Selai Tukunga, Faleola Mafi, Karen Fukofuka, Nick Wilson

## Abstract

**Introduction:** Fiscal policies are recommended as an effective policy to promote healthy nutrition and prevent non-communicable diseases. Sugar-sweetened beverage taxes are widely used, however there are fewer examples of taxes targeting unhealthy foods. Tonga introduced taxes targeting foods high in fat, salt, sugar from 2013 to 2018. This study examines the effect of these tax increases on food prices and import volumes.

**Methods:** Interrupted time series analysis using segmented generalised least squares regression was applied to assess the impacts of tax increases on retail prices and import volumes of taxed foods and their potential substitutes. Nine food tax increases were examined, varying from US$0.17/kg to US$1.08/kg, and averaging 28% of import value. Observed post-tax trends were compared with projected pre-tax trends (counterfactual), with adjustment for autocorrelation and potential time-varying effects by GDP, international visitor numbers, month, T$/US$ exchange rate, oil prices, and specific international food prices. Bootstrapping was used to estimate 95% confidence intervals, and random effects meta-analysis was used to summarise outcomes across food types.

**Results:** In the first-year post-tax, retail prices increased for 10 of the 11 taxed indicator foods, with an average increase of 13% (95%CI: 5% to 21%). Seven price increases were statistically significant: fresh and frozen chicken pieces (46% and 39%), mutton flaps (20%), corned beef (20%), ice cream (11%), salted beef (6%) and mayonnaise (3%). Import volumes decreased for eight of the nine taxed foods with an overall mean decrease of 41% (95%CI: 17% to 58%). Two import decreases were statistically significant: sausages (68% reduction) and ice cream (72%).

**Conclusion:** Food excise taxes in Tonga were associated with clear increases in retail prices and decreased imports of taxed foods. These findings strengthen the existing international evidence that excise taxes on unhealthy foods effectively raise prices and reduce supply.

**Summary box of key messages:** *What is already known on this topic:* In contrast with sugar-sweetened beverage taxes, only a handful of taxes on unhealthy food have been evaluated, with studies published from Denmark, Hungary, Mexico and some United States jurisdictions. These evaluations report small declines in purchasing or intake (eg, 3% to 5%), reflecting the low level of taxation (less than 10% of price). We could not identify any evaluations of food taxes outside of Europe or North America.

*What this study adds:* Tongan taxes targeted a range of unhealthy foods and were introduced at a relatively high level of taxation (averaging 28% of import price). Retail prices increased in 10 of 11 monitored foods, with an average price increase of 13% compared to counterfactual and a 68% pass-through. Import volumes decreased in 8 of 9 taxed foods with an average decrease of 41%. In the year after each tax was introduced government revenue increased by a total of US$2.8 million or US$28 per population.

*How this study might affect research, practice or policy:* This study strengthens the existing international evidence by demonstrating the effectiveness of unhealthy food excise taxes in raising prices and reducing the supply of unhealthy foods in a small island developing state. Study findings are supportive for policy makers considering introducing unhealthy food excise tax policies, to address high rates of non-communicable disease.

## Introduction

Fiscal policies such as taxes on unhealthy foods have been recommended by the World Health Organization to promote healthy nutrition and prevent non-communicable diseases (NCDs).(1) Overweight and nutrition-related factors have been reported as one of the largest causes of health loss,(2) premature deaths and health inequities.(3, 4) There is a direct association, for example, between exposure to ultra-processed food consumption and multiple health outcomes including mortality, cancer, cardiovascular and mental health.(5)

A growing number of jurisdictions have responded to increases in obesity and nutrition-related NCDs with fiscal policies. Sugar-sweetened beverage taxes are commonly used(6) and in a similar vein, a number of jurisdictions are adopting excise taxes on foods high in fat, salt and sugar (HFSS). There are 41 countries with examples of these taxes identified by the Global database on the Implementation of Food and Nutrition Action.(7) However, only a limited number of policies have been evaluated,(8) with observational studies examining HFSS food tax effects in Mexico, Hungary, Denmark and selected jurisdictions in the United States.(8, 9) Food tax levels were generally less than 10%, and tax impacts reflected this with small declines in purchasing or intake (eg, 5%,(10) 3%,(11) 4%(12)). No evaluations were available from Asia or the Pacific, Small Island Developing States or Low-Income Countries. Further HFSS tax research has been recommended for more informed policy making.(8, 9)

Tonga is a middle-income country in the South Pacific which has some of the highest obesity and diabetes rates in the world,(13) a modern phenomenon driven by globalisation and the availability of highly-processed energy-dense imported food. Tonga introduced excise taxes and tariffs from 2013 to 2018 that targeted foods HFSS. Tax increases were generally excise taxes, moderate to high levels, volumetric in nature (per kg) and they targeted a broad range of commonly consumed unhealthy food options, the vast majority of which are imported into the country. Tariff waivers on fish, fruit and vegetables were also introduced in 2013, 2015 and 2016 respectively. There was generally a low level of public awareness about fiscal policy changes (<20% population were aware about the 2017 tax changes).(6)

However, excise tax and tariff increases were expected to pass-through to prices and reduce the demand for taxed foods. On the other hand, taxing some HFSS foods and not others might increase the risk that consumption would shift towards other unhealthy food items that are not taxed (substitution).(14) Experts have recommended: broadening the tax base to include a broad range of HFSS foods, introducing a larger 20% to 50% level of tax increase,(15, 16) introducing waivers for healthy foods at the same time as excises, and improving the saliency (awareness) of the tax, with the expectation of improved effects.(9) Tonga’s tax policy aligns with several of these principles, presenting an opportunity to evaluate its real-world impact.

The aim of this study was therefore to quantify how Tongan food taxes affected food prices and import supply. Our focus was on foods that were taxed, with reporting also on potential substitute foods. It was expected that tax changes of over 10% will typically increase the shelf price of taxed foods; and that subsequently the importation volumes of taxed foods will decline, compared to pre-existing trends.

## Methods

For each food tax, an interrupted time series(17, 18) design was used to compare observed food prices and import volumes after the tax, with a ‘counterfactual’, that is the average expected projected outcomes with no tax change. An interrupted time series analysis is a commonly applied method for analysing natural experiments where the same population is compared before and after a population-level intervention. The method was selected based on its strength of design in reducing the risk of selection bias, time-varying confounding and suitability for assessing tax policy changes and population level outcomes.(17) Analysis was pre-planned and reported in a pre-published protocol.(19) Ethics approval was awarded by the Tonga National Health Ethics and Research Committee and the University of Otago Human Ethics Committee (D21/406), and research conformed to the principles embodied in the Declaration of Helsinki.

There was no formal patient or public involvement in this study. Preliminary findings were however presented at a meeting in Tonga to both local public and private stakeholders who gave interpretation and feedback. Study findings will be presented in conferences and in local meetings/media.

### Food excise taxes

Any excise or tariff targeting food was eligible for evaluation if it was introduced (or increased) between 2009 and 2019, the tax increase comprised more than 10% of import value, and there were outcome data for at least 12 months pre- and post-tax. The level of excise taxes (T$/kg) was compared to imported food value in the year of the tax change from (T$/kg) to give an ad valorem equivalent (AVE, %) tax rate.(20) This AVE enabled concurrent changes in excise and tariff to be combined (excluding consumption taxes), and was used to calculate tax elasticities (%change in import volume for each %change in tax). Tax elasticities were used to compare outcomes across different food tax rates and estimate the average impact of the food tax policies.

### Price data

Price data was provided by the Tonga Department of Statistics covering the period from September 2010 to December 2019 (ie, before the disruption associated with the Covid-19 pandemic in 2020). Monthly prices (inclusive of taxes) were collected from retail surveys for commonly consumed foods. The retail survey was redesigned in September 2017, thus interrupting the time series for some foods. Indicator foods were selected (Additional File: Table C) if they were subject to excise tax or were potential substitute foods, and they had a year of data available pre- and post-tax (uninterrupted by changes in survey method). Focus was given to prices collected in Tongatapu, the largest island with 74% of the population of Tonga, to ensure comparability in price data over time.

### Trade and revenue data

Customs datasets with import data were provided by the Tonga Ministry of Revenue & Customs for the period of January 2010 to December 2019. The dataset included date of payment, trade type (import/export), food type description, trade code (international Harmonised System [HS] of coding used for analysing trade flows), warehouse description, quantity (eg, kg or L), cost, insurance and freight value, and tax paid (tariff, excise, consumption tax, total in T$). The customs and revenue dataset also included rows for the revenue collected from local manufacturers of excise taxed foods. Identical duplicate rows were removed.

Import data was classified into food groups using the Harmonised codes described in the legislation for each tax (Additional File: Table B), and key words from the food type description were used for consistent categorisation of foods over time if pre-tax HS codes were not specific to the taxed food. Trade data quality was checked to identify any unusual missing data, trends and outliers. Data were cleaned using an automated process to recalculate outlying values from the warehouse description, with manual checks of remaining outliers (Additional File: Further Methods).

### Analysis

The primary study outcomes were (1) average price changes in selected indicator foods (T$ per item) and (2) import volumes into Tonga (kg per population) for taxed foods, in the first-year post-tax. Outcomes for potential substitute foods and outcomes two-years post-tax were also reported as secondary outcomes. Potential substitutes comprised foods that were not subject to tax increases or tax decreases or waivers during the study period. We report findings for potential substitute food if they could have replaced taxed foods in the usual consumption pattern and diet.

Interrupted time series analysis can be used to investigate the change in an underlying outcome trend and can control for other factors affecting this trend. Generalised least squares (GLS) regression was used to fit a segmented linear model to monthly data. A level and a trend term were included to model, the latter controlling for short-term fluctuations in the series and any secular trend.(21) Observed post-tax trends were compared with pre-tax trends projected forward. Testing and adjustment for autocorrelation and moving averages was informed by the unadjusted model using a graph of the residuals and a plot of the autocorrelation function and partial autocorrelation function. If the autocorrelation adjusted model was significantly different from the unadjusted model (log-likelihood ratio test), then the model with the best fit to the data was selected (lowest AIC). Testing and adjustment for autocorrelation and moving averages was informed by the unadjusted model using a graph of the residuals and a plot of the autocorrelation function and partial autocorrelation function. If the autocorrelation adjusted model was significantly different from the unadjusted model (log-likelihood ratio test), then the model with the best fit to the data was selected (lowest AIC).

The model was adjusted for potential (pre-selected) time-varying influences from factors that predict import volumes and prices of taxed foods. These were exchange rate (T$/US$), international oil prices (proxy for shipping costs), GDP, international visitor numbers, season, and international food prices (for chicken, mutton, beef, sugar and oranges if the outcome aligned with one of these products). The time-varying seasonal effect was modelled using the month of the data. If a more complex model including cyclones causing substantial damage in Tonga (eg, February 2018 Cyclone Gita, dummy variable) or price survey method change (September 2017, level and trend change) was a significantly better model fit, assessed using likelihood ratio tests, then these variables were also added to the model. The following equation provides an example of the fitted linear time series regression model for mutton flaps.

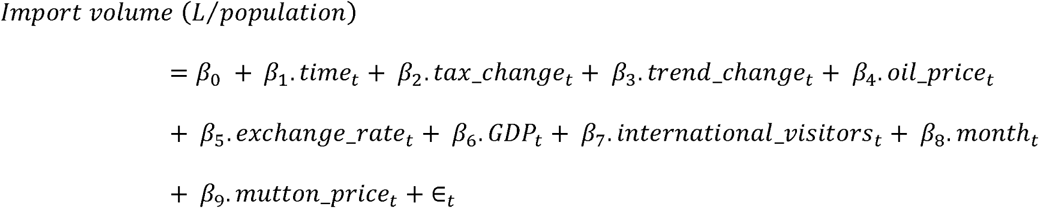

If there were multiple intervention points for the same food, these were evaluated within the same model. Both import volumes and prices were expected to only have positive values. The outcome values were log-transformed to predict non-negative average outcomes and stabilise variances. Multiplicative effects were estimated and interpreted as percentage changes and elasticities. Parametric multivariate simulation and bootstrapping were used to estimate 95% confidence intervals for the percentage change outcomes (using the coefficients and their standard errors from the GLS model and allowing for correlation between correlates). A random effects meta-analysis technique was used to pool effect sizes across different excised taxed foods to give the average tax effect. Tax pass-through was calculated as the modelled post-tax change in price compared to counterfactual (T$/kg) divided by the level of the tax in T$/L (Additional File: Table F).

Sensitivity analyses assessed the robustness of the key food price and trade results in pre-specified scenarios:

1. A two-month time lag between tax legislation and change in outcome was added to reflect the extra time it took for excise collection to begin after legislation was changed for some taxes.
2. Pre-tax data was limited to three years pre-tax change, to limit the influence of potential long-term historical trends.(17)

The change in government tax revenue was calculated from the summed excise, tariff and consumption tax revenue in the 12 months after each tax increase compared to the 12 months before.

All analyses were done using R version 4.3.1 [R Statistical Foundation, Vienna, AT]. GLS regression was fitted using the nlme package.(22)

## Results

From 2015 to 2018, Tonga implemented nine tax increases that were eligible for evaluation, of which seven were volumetric excise taxes and two were increases in ad valorem import tariffs. There was an average tax increase of 28%, as a proportion of a food’s import value (Table 1). Evaluated volumetric tax levels varied from T$0.40/kg (US$0.17) to T$2.50/kg (US$1.08) (Additional File: Table A).

**Table 1:** Impact of tax increases on taxed food prices and import volumes, for the first-year post-tax, Tonga

| Food product | Year | Tax ↑<br>(T\$/kg) | Tax ↑<br>(AVE%) | Average price<br>change (T\$/kg) | %Δ price | Import volume<br>change<br>(kg/person) | %Δ imports | Tax elasticity |
| --- | --- | --- | --- | --- | --- | --- | --- | --- |
| Turkey tails | 2015 | 0.48<sup>4</sup> | 15% | -0.34 (-1.22 to 0.53) | -6 (-18 to 11) | -1.4 (-6.5 to 0.6) | -46 (-79 to 40) | -3.1 (-5.3 to 2.7) |
|  | 2016 | 1.50 | 59% | -0.29 (-1.40 to 0.82) | -5 (-21 to 19) | -0.9 (-5.6 to 0.7) | -39 (-81 to 85) | -0.7 (-1.4 to 1.4) |
| Mutton<br>flaps | 2016 | 0.96<sup>4</sup> | 15% | **2.12 (1.25 to 2.99)** | **20 (11 to 30)** | -5.5 (-22.1 to 1.9) | -47 (-79 to 33) | -3.1 (-5.3 to 2.2) |
| Sausages | 2018 | 1.00 | 23% | 0.16 (-0.22 to 0.55) | 2 (-3 to 7) | **-8.1 (-24.0 to -1.2)** | **-68 (-87 to -21)** | **-3.0 (-3.8 to -0.9)** |
|  |  |  |  | 0.41 (-0.16 to 0.99)<sup>1</sup> | 18 (-6 to 59)<sup>1</sup> |  |  |  |
| Chicken leg<br>quarters | 2016 | 0.40 | 22% | **1.03 (0.80 to 1.27)<sup>5</sup>** | **39 (28 to 52)<sup>5</sup>** | 1.4 (-34.0 to 27.0) | 2 (-36 to 63) | 0.1 (-1.6 to 2.9) |
|  |  |  |  | **1.16 (0.82 to 1.50)<sup>6</sup>** | **46 (29 to 67)<sup>6</sup>** |  |  |  |
| Processed<br>beef | 2018 | 2.50 | 17% | **2.29 (2.01 to 2.56)<sup>2</sup>** | **20 (18 to 23)<sup>2</sup>** | -1.2 (-3.8 to 0.7) | -34 (-66 to 30) | -2.0 (-3.9 to 1.8) |
|  |  |  |  | **0.71 (0.13 to 1.29)<sup>3</sup>** | **6 (1 to 12)<sup>3</sup>** |  |  |  |
| Instant<br>noodles | 2016 | 2.00 | 20% | 0.06 (-0.03 to 0.16) | 6 (-3 to 18) | -7.4 (-33.4 to 0.8) | -60 (-88 to 17) | -3.0 (-4.4 to 0.9) |
| Mayonnaise | 2016 | 2.00 | 29% | **0.26 (0.00 to 0.53)** | **3 (0 to 6)** | 0.0 (-0.7 to 0.3) | -12 (-67 to 135) | -0.4 (-2.3 to 4.7) |
| Ice cream | 2016 | 1.50 | 48% | **1.46 (0.71 to 2.20)** | **11 (5 to 17)** | **-12.0 (-41.5 to -1.6)** | **-72 (-90 to -23)** | **-1.5 (-1.9 to -0.5)** |
| Overall<br>summary | 2015-2018 | 1.37 | 28% | - | **13 (5 to 21)** | - | **-41 (-58 to -17)** | **-1.5 (-2.1 to -0.6)** |

Notes: 1. Price change for hot dogs; 2. corned beef; 3. salted beef; 4. These were increases in import tariffs, not excise taxes like the remainder. Tariff increases were converted into T\$/kg using import unit value for the same year. 5. Price for frozen chicken pieces. 6. Price for fresh chicken pieces. AVE is the ad valorem size of the tax change as a percentage of import value. GLS segmented linear model adjusted for GDP, month, international visitor numbers, exchange rates and oil prices and autocorrelation effects if relevant. Overall summary presents meta-analysis random effects summary measures for the percentage change in price and imports. Tax elasticity is the percentage change in import volume divided by the tax increase (AVE%), and the summary measure of each in the case of the overall value. Bolded results were statistically significant, p<0.05.

### Taxed foods

#### Indicator food prices

Prices of taxed indicator foods all increased compared to the predicted counterfactual without tax, with turkey tails being the one exception (Table 1, Additional File: Figure A). The largest price increases were T$2.29/kg in the price of corned beef, T$2.12 for mutton flaps, and T$1.46/kg for ice cream (all statistically significant increases). The increases in indicator food prices were largest for fresh and frozen chicken pieces (46% and 39%), mutton flaps (20%), corned beef (20%), and ice cream (11%), with smaller increases for salted beef (6%) and mayonnaise (3%) (all significant). The 18% increase in the price of hot dogs had a confidence interval of -6% to 59%, suggesting a moderate but imprecise effect estimate. Price changes were small and not statistically significant for turkey tails (−6% in 2015 and - 5% in 2016), sausages (2%) and instant noodles (6%). Random effects meta-analysis summarised the price changes across taxed foods and found an overall average 13% (95%CI: 5% to 21%) increase in taxed indicator food prices, with a high level of heterogeneity (I^2^=95%, p<0.0001).

Tax pass-through followed a similar variability with an average pass-through 68% across the taxed indicator foods (Table F), and levels ranging from -71% to 258%. Under-shifting was most common, but there was significant over-shifting of the taxes on mutton flaps and chicken leg quarters.

#### Import volumes

Compared to the counterfactuals, there was a decrease in import volumes (percentage change) after eight of the nine tax increases (Table 1, Additional File: Figure A), with chicken leg quarters the only exception (2% non-significant increase). The largest decreases in imports were for sausages (−68%, 95%CI: -87 to -21%) and ice cream (−72%, CI: -90 to - 23%). Meta-analysis revealed an average decrease of 41% (CI: 17% to 58%) in import volumes across the nine food tax changes (moderate heterogeneity I^2^=30%). For taxes where two years of follow-up was possible, the changes in import volumes were very similar to the changes in the first year (Additional File: Figure B).

Tax elasticities of import volumes varied from 0.1 for chicken leg quarters to -3.1 for mutton flaps, with sausages, instant noodles and turkey tails (in 2015) also at -3.0 (Table 1). The average tax elasticity (calculated from the meta-analysis on import volume changes) was approximately -1.5 (CI: -2.1 to -0.6), which is a 1.5% average decrease in imports for every 1% increase in tax (as a proportion of import value) and indicative that import volumes were elastic to tax changes.

Post-tax price increases corresponded to post-tax decreases in import volumes (as a percentage change) for mutton flaps, sausages, processed beef, instant noodles, mayonnaise and ice cream. Chicken leg quarters were a notable exception with large price increases but no evidence of change in import volumes. The pattern for turkey tails also varied, where even though there were increases in tax and decreased imports, the corresponding turkey tail indicator price declined.

#### Local food manufacturing

Of the three excise taxed foods that were also locally produced in Tonga as well as being imported, only sausages (taxed at half the level of imports), may have had increased local production. The tax revenue collected on locally produced sausages had an increasing post-tax trend, but initially incomplete revenue collection may underestimate early volumes. Excise taxed locally produced sausages, made up 13% of the total supply of excise taxed sausages by volume in the 2019 calendar year. The equivalent level of market share by volume for locally produced ice cream was 2.1% in 2019, and 0.03% for instant noodles.

#### Tax revenue to government

Generally consistent with tax increases, there were large increases in total food tax revenue in the year following each of the tax changes, compared to the year prior. The exception was for instant noodles, which at the same time had a large decline in imported volume (Additional File: Table E). The total revenue increase on imported products was T$6,576,218 (US$2,791,276, or US$28 per person).

Revenue changes, as a percentage of import values, approximated the level of the tax increases with two main exceptions. The revenue increases were more than the tax increases for the 2016 turkey tail excise (99% revenue vs 59% tax) and for the 2016 mayonnaise excise (45% revenue vs 29% tax).

## Sensitivity analyses

Sensitivity analyses are reported in Table 2 and show a relatively similar pattern of results for a shorter pre-trend timeframe and when a two-month transition period was allowed for in the model. Revenue was not collected until September after the July introduction of excise taxes on turkey tails, chicken leg quarters and ice cream in 2016 and processed beef in 2018. The impact of modelling a two-month time lag on the findings for these foods was mixed, and did not impact overall study conclusions.

**Table 2:** Sensitivity analyses of tax increases on taxed import volumes, for the first-year post-tax, Tonga

| Food product | Year | Original analysis | Three-years pre-tax trend | Adjusting for a two-month time lag |
| --- | --- | --- | --- | --- |
| turkey tails | 2015<sup>1</sup> | -46 (-79 to 40) | **-85 (-97 to -30)** | na |
|  | 2016 | -39 (-81 to 85) | -12 (-84 to 311) | -1 (-88 to 512) |
| mutton flaps | 2016<sup>1</sup> | -47 (-79 to 33) | -45 (-85 to 98) | -32 (-75 to 84) |
| chicken leg quarters | 2016 | 2 (-36 to 63) | -19 (-60 to 62) | -5 (-35 to 38) |
| sausages | 2018 | **-68 (-87 to -21)** | **-71 (-90 to -17)** | **-68 (-88 to -16)** |
| processed beef | 2018 | -34 (-66 to 30) | -57 (-85 to 19) | -44 (-72 to 12) |
| instant noodles | 2016 | -60 (-88 to 17) | -58 (-89 to 40) | -67 (-91 to 12) |
| mayonnaise | 2016 | -12 (-67 to 135) | -22 (-78 to 176) | 5 (-62 to 196) |
| ice cream | 2016 | **-72 (-90 to -23)** | -64 (-91 to 38) | **-67 (-88 to -7)** |

Notes: 1. These were increases in import tariffs, not excise taxes like the other tax changes. GLS segmented linear model adjusted for GDP, month, international visitor numbers, exchange rates and oil prices and autocorrelation effects if relevant.

### Untaxed potential substitute foods

Changes in import volumes of untaxed meat (such as chicken, beef, pork and lamb, but not fish) were examined after increased taxes on turkey tails, mutton flaps and chicken leg quarters (July 2016), and sausage and corned beef taxes (July 2018). In 2016, import volumes of untaxed meat increased by 115% (CI: -17% to 459%) consistent with a substitution effect, but in 2018 untaxed meat import volumes decreased by 46% (CI: -80% to 46%, the same time tax was waivered on chicken leg quarters). The corresponding decreases in the price of whole frozen chicken and lamb chops in 2016 may have contributed to the suggested substitution effect.

There was no evidence of any substitution to imported and untaxed types of crisps and snacks, sugar, sauces, or other processed foods (including pasta), after taxes were increased on instant noodles, ice cream and mayonnaise in July 2016. This was despite decreases in the price of potential substitute products such as bongos (a type of snack in the crisps and snacks group). Conversely, a significant increase in price of sugar may have contributed to the apparent lack of substitution. (Additional File: Figures C and D)

## Discussion

### Summary and comparisons with the international literature

#### Taxed foods

Study findings document further evidence that taxes targeting unhealthy food high in fat, salt, sugar in Tonga from 2015 to 2018 were associated with significant increases in taxed food prices and reduced import supply of these foods. Given the almost exclusive supply of these foods by international trade, decreased imports suggest decreased levels of sales and purchasing of taxed foods in Tonga. For example, among taxed foods, only locally produced sausages (taxed at half the level of imports), had a significant market share, reaching 13% of the total excise taxed sausage market by volume in 2019.

The average tax level was 28% of the import value for the foods examined in this study. In response, the average corresponding retail price increases were 13% (CI: 5% to 21%) and the average tax pass-through was 68%. At the same time, there was a large drop in import volumes averaging 41% (CI: 17% to 58%). Even with the decline in imports, there remained a substantial increase in tax revenue for the Tongan Government in the first year after each tax, totalling T$6,576,218 (US$2,791,276), or T$65 per resident.

The average HFSS food tax pass-through in Tonga was equivalent to average pass-through rates for sugar-sweetened beverage taxes reported in the literature, eg, 67% and 70% in meta-analyses,(23, 24) albeit that there was substantial variation eg, by study, food type, and levels of competition,(25) similar to what was seen across food taxes in Tonga.

Some foods appeared to be more responsive to taxation. Tax increases on turkey tails (in 2015, tax elasticity -3.1), mutton flaps (−3.1), sausages (−3.0), processed beef (−2.0), instant noodles (−3.0), and ice cream (−1.5), appeared to be particularly successful in reducing import supply, although results were only statistically significant for ice cream. The high declines in import volumes (46%, 47%, 68%, 34%, 60% and 72%) were associated with moderate to high tax increases respectively (15%, 15%, 23%, 17%, 20%, and 48%), indicating elastic demand. The corresponding indicator foods all increased in price, at least a little (except for turkey tails), and half were statistically significant increases. It may be that these food groups comprise more ‘luxury’ foods that are therefore more sensitive to price changes than staple foods.(26, 27)

There are other reasons the elasticities reported in this study are generally larger than those seen elsewhere.(26) The tax increases in Tonga were moderate to large compared to elsewhere, and consumers in this middle-income country may be relatively more price sensitive.(26) The availability and supply of cheaper local food options, may have improved the ability to switch away from imported to locally sourced foods. There also may be a potential “Giffen effect”(28) whereby taxes on luxury foods result in reduced consumption of more basic foods, due to increased food budget constraints. Budget constraints may help to explain the reduced spending on many imported foods around the time taxes were introduced.

Foods with least sensitive tax elasticities were chicken leg quarters (0.1), mayonnaise (−0.4), and turkey tails (in 2016, -0.7). There were very small changes in price for mayonnaise (3%) and turkey tails (−5%) with a low tax pass-through. It is also possible that there is less price sensitivity for low volume condiments like mayonnaise that consumers may typically use sparingly, or if the product is complementary to another food like mayonnaise and potatoes in potato salad. The low tax elasticity for chicken leg quarters, occurred despite large statistically significant price increases of 39% and 46%. Chicken leg quarters are one of the cheapest forms of protein in Tonga and remained a staple in the diet post-tax(6) contributing to the low tax elasticity.(26) The excise on chicken leg quarters was subsequently reversed in 2018, not least because low-income households continued to purchase them increasing the likely regressive nature of the tax on this food.(6)

The World Bank report(6) gives price elasticities after four of the 2016 Tonga food tax increases: -1.9 for mutton flaps, -1.4 for ice cream (both statistically significant), -0.4 for chicken leg quarters and -0.4 for instant noodles (albeit the latter two not statistically significant).(6) Our tax elasticity findings for ice cream (−1.5) and chicken leg quarters (0.1) were approximately consistent with this, however instant noodles (tax elasticity of -3.0) and mutton flaps (−3.1) in our analysis were associated with relatively greater declines in import volumes (likely due to adjustment for pre-existing trends and time-varying factors).

In the Tonga setting, the price signal which discourages purchasing appears to be a major pathway for how food taxes may be working. Taxes may also act via public awareness about which foods are taxed, industry reformulation of foods, and increased availability of healthier options. However, public awareness was low in Tonga and there was limited ability for industry to avoid taxation (eg via reformulation) given the nature of the tax design being based on volume of products and Tonga being a small market. The availability of healthier options is considered to remain a major challenge,(6) but we note that Tonga has a local seafood industry and relatively low-priced locally produced vegetables and fruits (with home gardens being common).

#### Substitution

We were interested in whether consumers switched from taxed imported foods to cheaper and/or other less healthy products. There was a suggestion of substitution to untaxed imported meat after the 2016 taxes on mutton flaps, turkey tails, and chicken leg quarters. In 2016, there was also an increasing trend in imports of sausages, after the import tariff on sausages was removed. However, there was no evidence of untaxed meat substitution in 2018 when the taxes were broadened to sausages and processed beef, though imports of chicken leg quarters did increase after they were exempted from excise tax in the same legislation change. We found no evidence of substitution to untaxed imported foods like snacks and crisps that also may be HFSS.

Participants in interviews and focus groups for the World Bank study reported switching to substitutes like corned beef and locally manufactured instant noodles and ice cream after the 2016-tax changes.(6) This is consistent with the increasing trend in processed beef import volumes from July 2016 (Additional File: Figure A). But we did not identify any large shift to locally manufactured products, although we could not rule it out given potentially incomplete revenue collection.(6)

The decline in import volumes of untaxed imported foods compared to the counterfactuals in both 2016 and 2018 suggests that consumers may have been obtaining more food from local sources not subject to excise, including purchasing locally grown foods, and growing their own food (eg, taro, yams, bananas, coconuts, and fish). Other locally sourced food includes bread and rolls, and other baked goods. Retail price trends for common locally sourced foods (such as seafood, vegetables and eggs) were mixed (Additional File: Figure E).

### Study strengths and limitations

This, interrupted time series analysis is a natural experiment design evaluating a food excise tax strategy implemented in Tonga from 2015 to 2018. Advantages of studying these taxes in Tonga are: (i) the relatively high level and breadth of the food taxes; and (ii) as an island nation the imported food is relatively well-measured as it comes via cargo shipping (no land-border trade). The analysis pieces together historical trade and statistical data to compare pre- and post-tax trends in price and import volumes adjusted for time-varying factors and directly estimates tax pass-through to consumers. Prices and import volumes are proximal outcomes to taxes and so are less prone to errors or bias than more distal outcomes or longer-term outcomes. The research team (which included a Tongan doctor), met with local stakeholders to understand contextual factors that may have influenced study findings. Meta-analyses were used to overcome the limited power in many of the single food analyses and to estimate an overall effect of tax change across food types.

There were potential limitations. The import dataset was affected by changes in coding over time, and changes in the quality of data entry. This was addressed by using key word searches, automated checks to ensure consistent units, and excluding poor quality data (eg, confectionery imports). If measurement error remains and occurs randomly, it is likely to have a conservative impact on the reported results. Model specifications can affect results; however, sensitivity analyses were reassuring. Occasionally counterfactuals were influenced by the most recent data points, eg, the modelled counterfactual for sausage imports after the 2018 tax was influenced by the upward 2017 to 2018 pre-tax trend, potentially overestimating the decline in imports. To avoid any bias from changing model specifications we stuck closely to the pre-planned analysis.

Time-varying confounding is difficult to completely and precisely adjust for. International food prices were not available for every model. There may have been public promotion of purchasing local food or growing food that promoted a move away from imports. Price controls may have reduced the price changes in some imported goods.

The AVE% calculation of tax level is expected to overestimate tax size as a proportion of retail prices, because the import unit value is lower than retail price. Therefore, the reported tax elasticity may underestimate true effects.

We did not have adequate data with which to assess pre- and post-tax trends in consumption of locally sourced foods and manufacturing . Future research is planned to assess the impact of tax waivers on prices and import volumes, and heterogeneity of purchasing outcomes by household income.

### Potential implications for food tax policy design

These findings add to the existing international evidence(10–12) that food excise taxes can be effective in raising prices and reducing supply of unhealthy foods in different settings.

This is particularly true in this middle-income country where the tax impact has largely occurred via price changes, and tax elasticities appear to be relatively high (−1.5 on average). There was no major shift to other unhealthy imports after these broad-based taxes were introduced, except for the suggested increase in untaxed meat imports when turkey tails, mutton flaps and chicken leg quarters were taxed in 2016.

To improve food tax policies, the scope of included products in excise taxes on food HFSS could be broadened to include a wide range of commonly consumed HFSS and energy dense food products. In Tonga, for example, this may include processed foods and sugars that are not currently subject to excise taxes.

Inbuilt mechanisms to allow routine annual adjustments for inflation would ensure the real value of the taxes is maintained, sustaining their effectiveness in discouraging unhealthy food consumption. Without adjustment for inflation the impact of excise taxes is likely to decline over time in Tonga.

Revenue from excise taxes can be a stable source of government income, useful for investing in health and supporting local food production. A dedicated proportion of the tax revenue could be used to fund the health system, health and equity initiatives and local farmers to increase food production. This could include improving access, availability and affordability of local fruit and vegetables as well as local fish and seafood,. The availability and affordability of healthy foods has been cited as a major issue in Tonga.(6)

Public awareness campaigns are recommended to inform the population about the rationale and benefits of HFSS food taxes, emphasising their role in improving health outcomes and supporting local food sector development. This would further help to build and sustain public acceptance of fiscal measures.

Finally, tax design refinements could include targeting the levels of fat, salt or sugar in taxed foods, to more precisely address the unhealthiest options. This would incentivise better quality imported food and may motivate reformulation(29) of locally manufactured products. The sugar-sweetened beverage excise in Tonga for example is levied based on thresholds of sugar content. Nutrient profiling tools, such as that produced for Tonga,(30) can be used to precisely categorise which foods are taxed and regulated, such as limits on which places foods can be sold eg, in schools. Verification requirements on nutrient level policies requires resourcing. Similarly, implementation of excise collection from local manufacturers requires resourcing for improved nutrition outcomes, and equal levels of excise taxes on imported and locally manufactured foods would support this.

## Conclusion

HFSS food excise taxes in Tonga were associated with increased prices and decreased imports of taxed foods. Findings strengthen the existing international evidence that food excise can be effective in raising prices and reducing the supply of unhealthy foods. Tongan analyses extend the number of settings where HFSS taxes have been evaluated and found to have positive effects. These policies are likely to lead to improved population nutrition and a lower burden of non-communicable diseases. Study findings should encourage policy makers to adopt and refine HFSS food tax policies.

## Declarations

### Availability of data and materials

The data that support the findings of this study are available from Tonga Ministry of Revenue & Customs and the Tonga Department of Statistics. Ethical approval was required for their use. Data are however available from the authors upon reasonable request and with permission of the Tonga Ministry of Revenue & Customs and the Tonga Department of Statistics.

### Competing interests

None declared.

## Funding

Marsden Project from Royal Society 23-UOO-157. University of Otago Research Grant 2022. The funder had no role in the design of the study and collection, analysis, and interpretation of data and in writing the manuscript.

### Authors’ contributions

AT, VP, AK and NW conceptualised the study and obtained funding. ST and FM extracted the datasets and supported data interpretation. AT and AK did the data analysis. AT prepared the tables and figures and wrote the first draft of the manuscript. AT, VP and NW investigated key contextual factors with Tongan experts. KF supported dissemination of preliminary results. All authors’ read, reviewed and edited the manuscript and approved the final version.

## Acknowledgements

Tonga Ministry of Revenue & Customs provided the trade dataset and expertise. Tonga Department of Statistics provided data on population projections, GDP, international visitor numbers, for differing time periods. Thank you to Dr Si Thu Win Tin (Pacific Community), Dr Viliami Manu (Ministry of Agriculture), Feleti Lynch (Ministry of Revenue & Customs), and Sandra Fifita (Ministry of Trade and Economic Development) for comments on the final draft. Special thanks to Tutulu Finau for assistance with price data, who has sadly passed.

## Author reflexivity statement

Tongan stakeholders met with the study team and supported the conceptualisation of the study research question, expressing their interest in the overall impact of excise tax policies on the healthiness of import foods, which is also reflected in Tonga’s non-communicable disease plan. Early career researchers who collated the data were funded for their time and included as co-authors, and collaborative meetings were used to discuss data quality improvements and offer further input on data cleaning. Preliminary analyses were shared in person at a key stakeholder meeting in Tonga in July 2024 and audience feedback at the meeting (and subsequently) informed improvements in the manuscript. Our team maintains relationships with key stakeholders and collaborators in Tonga which includes contributing to key Tongan non-communicable disease control meetings and health promotion training. The study team has trained Tongan nursing staff on research methods. We continue to use research funding to support Tongan colleagues, for example in drafting a policy brief. A New Zealand based Tongan doctor was a key member of our small study team facilitating our relationships in Tonga.

## Figure titles and legends

**Figure 1:**
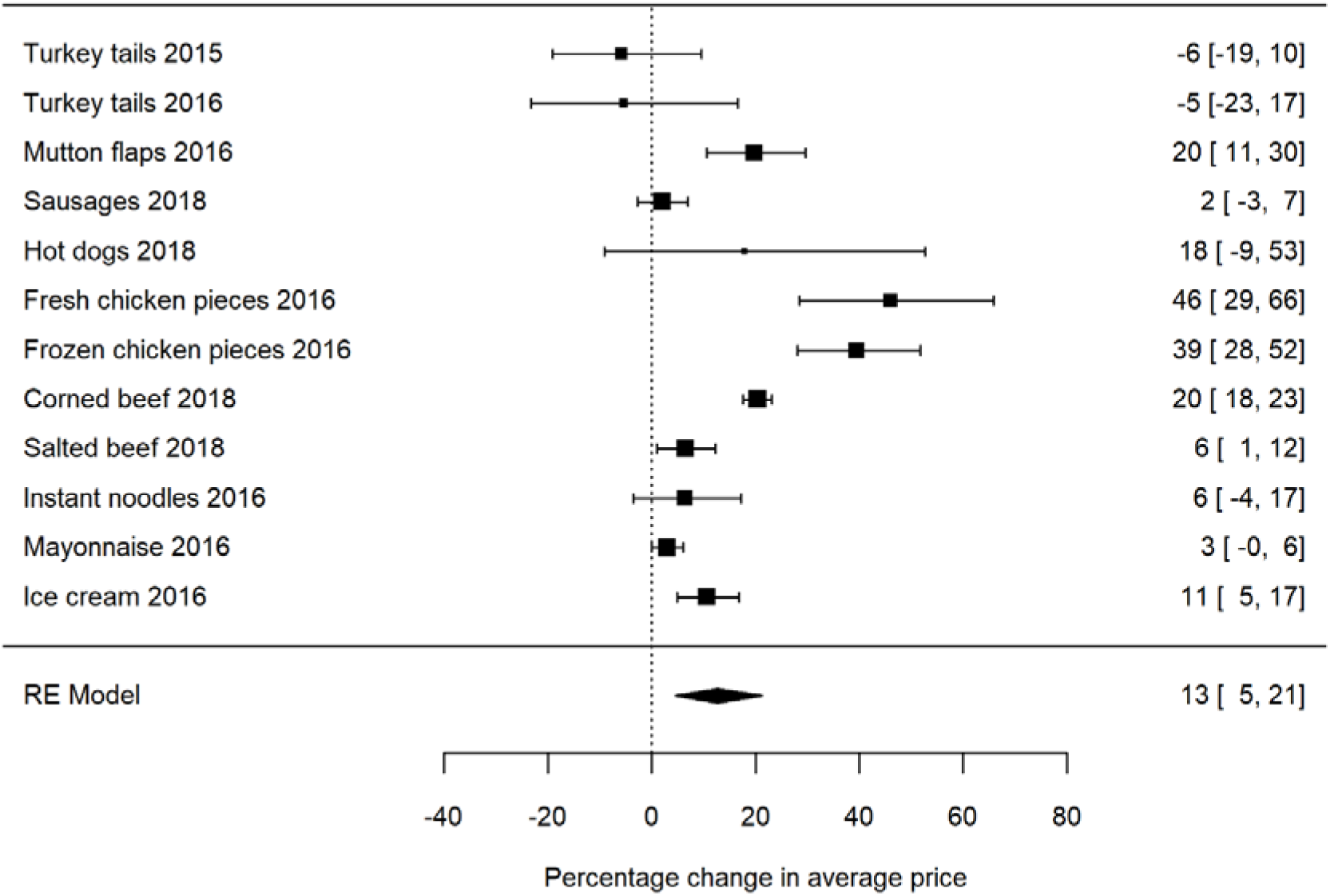
Meta-analysis summarising excise tax effects on taxed food retail prices in the first-year post-tax. Note: Data sourced from Tonga Department of Statistics.

**Figure 2:**
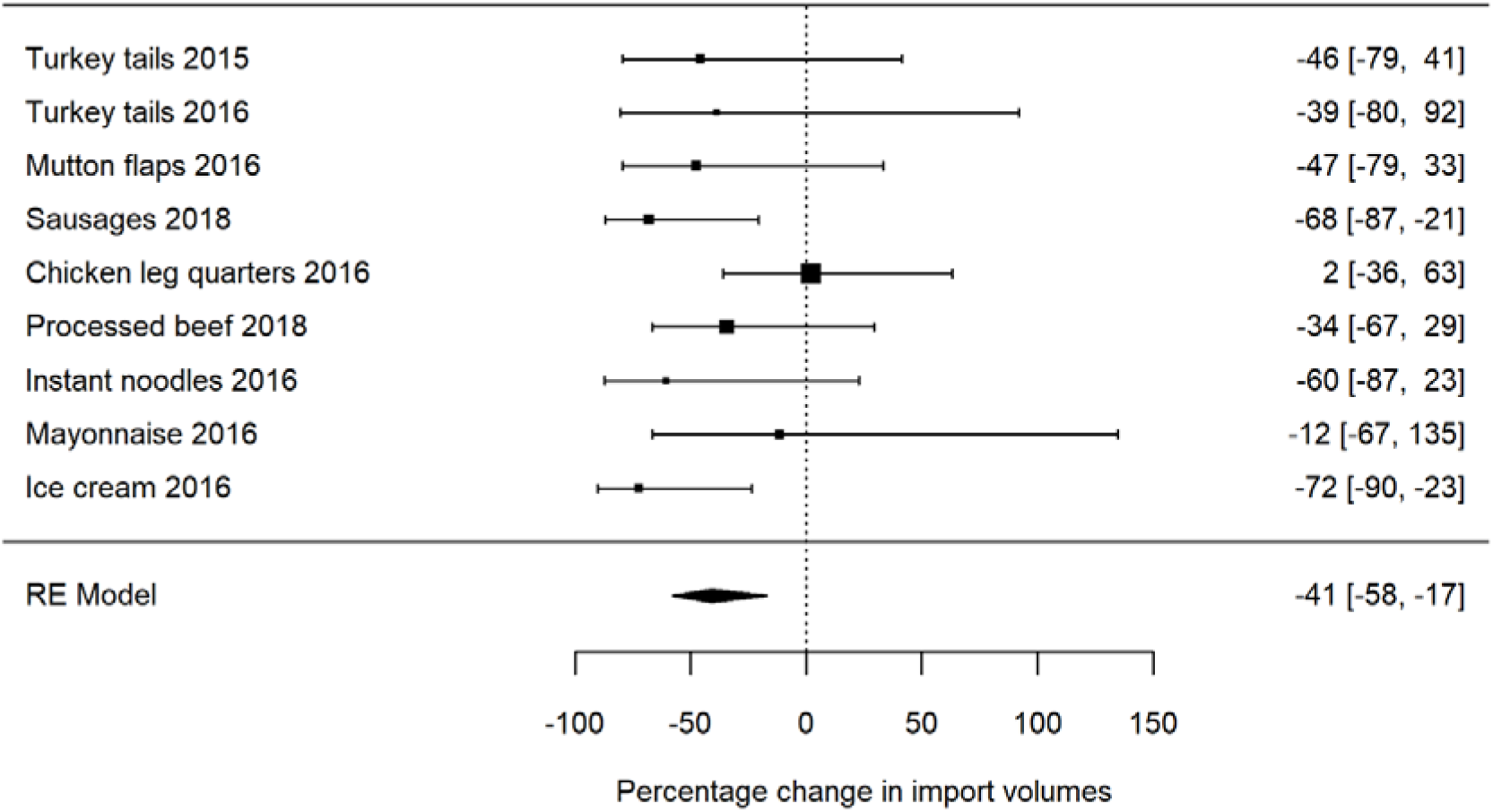
Meta-analysis summarising excise tax effects on taxed food import volumes in the first-year post-tax. Note: Data sourced from Tonga Ministry of Revenue & Customs.

## Additional file

Supplementary material is available from Additional file.docx. Further methods information describes data cleaning, the excise changes, and coding of food items. Additional results are reported including interrupted time series figures, two-year outcomes, revenue changes, pass-through rates and potential substitute food outcomes.

## Notes

### Competing Interest Statement

The authors have declared no competing interest.

### Clinical Protocols

http://hdl.handle.net/10523/166222024

### Author Declarations

Ethics approval was awarded by the Tonga National Health Ethics and Research Committee and the University of Otago Human Ethics Committee (D21/406)

