## Additional File for "Impact of Tongan taxes targeting foods high in fat, salt and sugar: Interrupted time series analyses"

### Further methods

#### Trade data quality checks and cleaning

Food categories (eg, for taxed foods in 2012 and 2019) were checked by comparing the number of rows identified by text fields for a given product with the number of rows identified by HS codes (eg, specified in the tax policy). This helped to identify any older HS codes specific to the food in question and to then include them. If 2012 HS codes were not specific to the food in interest, but a number of food products were coded in broad HS categories, then unique text fields were included to identify the food products across time. This was done for turkey tails, chicken leg quarters, mutton flaps, processed beef, noodles and mayonnaise. Text fields were checked to ensure no incidental foods were added, eg, at the <1% level. In some cases, HS codes have been adjusted over time with more specific categories introduced, including when taxes were implemented. In these cases, the most consistent method was to use text fields to identify the imported product over time and not use HS codes (eg, for mayonnaise).

Revenue was also assessed for the impact of interventions on government tax revenue in simple plots. This allowed us to check the timing of tax changes, taxed food coding, and the coverage of implementation of the excise taxes and tax waivers on foods.

**Missing data**

In the customs dataset, the level of missingness for quantity and warehouse description was assessed by food type and year. In 2011, there was a higher level of missing volume (1.7% of food import entries) and warehouse description (61%), than in other years. By food, the highest levels of missing volume data were for sauces (3.2%) and mayonnaise (1.1%) with less than or equal to 0.1% missing for all other foods. The proportion of quantity entries equal to one were also assessed, as this was sometimes used as a default entry. The proportion of quantity entries equal to one was highest in 2019 (10.7%) followed by 2018 (6.5%), 2017 (4.7%) and 2011 (3.7%). By food, it was highest for ham (44%), mayonnaise (29%), legumes and nuts (16%), and confectionery (12%).

Outcome trends by food product were observed to check for any obvious coding errors or missing data. This is particularly important for checking the completeness of the extracted data for import volumes, and understanding the impact of methods changes in the price datasets.

**Range checks**

Very high peaks in quantity of food imports indicated possible errors in the quantities recorded in the import dataset, and this was investigated by comparing the warehouse description field with quantity (‘HS Qty’) and units. Many errors appeared to arise from the use of different units ie, grams (42% of warehouse descriptions) or pounds rather than kilograms, and gallons or millilitres rather than litres. For example, there were many errors to the factor of one thousand, where grams were confused with kg in calculating volumes. If a high level of errors was identified and this was unable to be corrected, a food was subsequently excluded from the analysis (eg, confectionery).

An automated process was set-up to identify and process customs entries where:

- quantities were two or more standard deviations greater than the mean for the food group (because the largest values are likely to have the biggest impact on import trends),
- the ratio of CIF value to volume was two or more standard deviations greater than the mean for the food group, and
- where volume was equal to one (and may have been mis-entered).

For these entries, the numbers from the warehouse description (if it was not missing and reported at least one unit) were multiplied to calculate volume in kilograms, using adjustors for any units that were given. Automated volumes were applied to about 2.9% of customs entries. This process was not applied if there were:

- any unusual characters in the warehouse description such as “[,=,/,(,-,],%”, which may indicate that it would be inappropriate to multiply all numbers in the text field.
- more than five separate numbers reported in the warehouse description.

The checking process was used to refine the algorithm, identify the level of any remaining errors, and correct them. We checked and manually corrected any errors in:

- the 10 highest volumes for each food that were auto replaced, to check whether the cleaning logarithm was working.
- the 10 highest volumes for each food that were not replaced by the logarithm, to check for any potentially meaningful errors that were not picked up by the cleaning process.
- all volumes that were corrected and replaced, using four or more numbers extracted from the warehouse description (given the greater likelihood of errors in this group).
- all volumes that were more than 2 standard deviations from the mean (quantity and value per volume) that had miscellaneous characters (not automatically replaced), to fix any remaining errors.

#### Table A: Food excise tax changes in Tonga, 2010-2019

| **Food product** | **HS codes** | **Date of excise change**  (month excise started to be collected) | **Excise level** | **Date of tariff change (month tariff collection changed)** | **Tariff changes** | **Notes** |
| --- | --- | --- | --- | --- | --- | --- |
| animal fat (eg, lard) | 1501.00.00  1501.10.00  1501.20.00  1501.90.00  1502.00.00  1502.10.00  1502.90.00 | 13 Aug 2013 (Aug) | T$1/kg | 2013 (Jul) | 15% to 0% | Too many zero counts to be able to model |
|  |  | 1 Jul 2016 | T$2/kg |  | - |  |
| turkey tails | 0207.26.10  0207.27.10 | 3 Jul 2015 | - | 2015 (Jul) | 0% to 15% |  |
|  |  | 1 Jul 2016 (Sep) | T$1.50/kg |  | (stayed at 15%) |  |
|  |  | 9 Jul 2018 | T$2/kg | 2018 (Jun)* | 15% to 0% | The excise and tariff change resulted in a <10% tax increase |
| chicken leg quarter cuts (reversed in 2018) | 0207.13.10  0207.14.10 | 1 Jul 2016 (Sep) | T$0.40/kg |  | (stayed at 0%) |  |
| mutton flaps | 0204.22.10  0204.22.20  0204.42.10  0204.42.20 | 1 Jul 2016 | - | 2016 (Aug) | 0% to 15% |  |
|  |  | 1 Jul 2017 (Aug) | T$1.15/kg | 2017 (Jun) | 15% to 0% | The excise and tariff change resulted in a <10% tax increase |
|  |  | 1 Jul 2020 | banned import of all meat of mutton and lamb breast(26) |  |  |  |
| sausages | 1601.00.19  1601.00.29  1601.00.99 | 1 Jul 2016 | - | 2016 (Sep) | 20% to 0% | Includes tariff removal in 2016 and then excise increase in 2018 |
|  |  | 9 Jul 2018 (Jul) | T$1/kg |  | - |  |
| processed beef eg, corned beef | 1602.50.00 | 9 Jul 2018 (Sep) | T$2.50/kg |  | (stayed at 0%) |  |
| instant noodles (27) | 1902.19.10 | 3 Jul 2015 (Jul) | T$1/kg | 2015 (Aug) | 15% to 0% | The excise and tariff change resulted in a <10% tax increase |
|  |  | 1 Jul 2016 | T$2/kg |  | - |  |
| mayonnaise | 2103.90.10  2103.90.90 | 1 Jul 2016 (Aug) | T$2/kg | 2016 (tariff revenue continued to be collected) | (stayed at 15%) |  |
| ice cream | 2105.00.00  2105.00.90 | 1 Jul 2016 (Sep) | T$1.50/L |  | (stayed at 15%) |  |
| sugar confectionery, chewing gum & chocolate | 1704.10.00  1704.90.00  1806.20.00  1806.31.00  1806.32.00  1806.90.00 | 1 Jul 2017 (Jul) | T$5/kg | 2017 (Jul) | 20% to 0%  (15% to 0% for 1704.90) | Too much poor-quality import data to model accurately, from incorrect units. Also tariff waived. |
| sweet biscuits, waffles, wafers | 1905.31.00  1905.32.00 | 1 Jul 2017 (Aug) | T$1.50/kg | 2017 (Jul) | 15% to 0% (stayed at 0% 1905.32 for wafers and waffles) | Excise replaced the tariff, so <10% increase in tax |

Note: * Tariff change based on change in revenue data only. T$ is Tonga Pa’anga, equivalent to US$0.43 at the time of analysis. Tariff waivers on imported food were also introduced in 2013 on fresh and tinned fish, vegetable oils; and in 2015 on fresh and frozen vegetables and fresh fruit. Source data: Tonga Ministry of Revenue & Customs.(28-32)

#### Table B: Identification of food items subject to excise tax or import tariff increases in the Customs dataset

| **Category** | **Food product** | **HS codes (and abbreviated/misspelt descriptive terms)** |
| --- | --- | --- |
| Taxed meat & fat | turkey tails | 0207.24.00  0207.26.10  0207.27.10  In description:  ‘turkey tail’  ‘turket tail’ |
|  | chicken leg quarter cuts (reversed in 2018) | In description:  ‘Chicken Legs Quarter’  ‘Chicken Leg Quarter’  ‘chicken leg q’  'Chicken Leq Quarter'  ‘Chicken leg 1/4’  'Chiken Leg Quarter'  'Chicken Leq Quarter'  'Chicken LQ'  'Chicken Thigh Quarter'  'Chicken Qleg'  'Chix Leg Qty'  ‘Chicjen Leg Qrts’  ‘Chick leg quarter’  ‘Chic leg quarter’  (0207.13.10 and 0207.14.10 were excluded because they did not exist pre-tax) |
|  | mutton flaps | 0204.22.10  0204.22.20  0204.42.10  0204.42.20  0204.22.30  0204.22.40  0204.42.30  0204.42.40  ‘flap’ |
|  | sausages | 1601.00.10  1601.00.90  1601.00.19  1601.00.29  1601.00.99 |
|  | sausages manufactured in Tonga (post tax only) | 1601.00.11 (2018 onwards only)  1601.00.21  1601.00.91 |
|  | processed beef eg, corned beef | 1602.50.00  ‘corned beef’  ‘c/beef’  ‘c.beef’ |
| Taxed other | instant noodles | 1902.19.00  1902.19.10  1902.19.19  ‘noodle’ |
|  | instant noodles manufactured in Tonga (post tax only) | 1902.19.11 |
|  | mayonnaise | In description:  ‘mayo’  'Myannaise'  'Mayuonaise'  'Myonaise'  'm,ayonnasie'  'myaonnaise'  (2103.90.10 was not used because it did not exist pre-tax and mayo was in a broad category with other sauces) |
|  | ice cream | 2105.00.90  2105.00.00 |
|  | ice cream manufactured in Tonga (post tax only) | 2105.00.10 |

#### Table C: Indicator food products used in the price analysis, including taxed foods and potential substitute products

|  | **Tonga food prices** | **Pre-tax period** | **Post-tax period(s)** | **Code(s)** |
| --- | --- | --- | --- | --- |
| **Taxed foods** | Noodles, 2 minute / Noodles | 2010-15 | 2015-16  2016-23 | AD19, ZZ7 |
|  | Turkey tails | 2010-15 | 2015-16  2016-18  2018-23 | AB48, ZZ16 |
|  | Mutton flaps, frozen | 2010-16 | 2016-17  2017-23 | AB12, ZZ13 |
|  | Chicken pieces, frozen | 2010-16 | 2016-17 | AB37 |
|  | Fresh Chicken Pieces | 2010-16 | 2016-17 | AB46 |
|  | Saveloys or hot dogs, pkt / Hot dogs | 2010-16 | 2016-18  2018-23 | AB18, ZZ17 |
|  | Saveloys | 2010-16 | 2016-18 | AB43 |
|  | Sausages, (local) / Sausages | 2010-16 | 2016-18  2018-23 | AB20, ZZ18 |
|  | Tinned corned beef | 2010-18 | 2018-23 | AB14, ZZ19 |
|  | Salted Beef | 2010-18 | 2018-23 | AB47, ZZ11 |
|  | Salad dressing (946ml-Best Food) / Mayonnaise | 2010-16 | 2016-23 | AE29, ZZ37 |
|  | Ice Cream, Imported / ice cream bowl | 2010-16 | 2016-23 | AE44, ZZ33 |
| **Untaxed meat** | Beef, rump steak | 2010-17 | na | AB11 |
|  | Chicken, whole frozen | 2010-17 | na | AB36 |
|  | Lamb chops | 2010-17 | na | AB49 |
|  | Meat pie (fresh / frozen) | 2010-17 | na | AB50 |
|  | Other Beef | 2010-17 | na | AB52 |
|  | Suckling pig | 2010-23 | na | AB22, ZZ46 |
| **Other staples** | Cabin bread / Breakfast crackers | 2010-23 | na | AD13, ZZ5 |
|  | Weetbix | 2010-23 | na | AD19, ZZ8 |
|  | Flour, plain, loose | 2010-23 | na | AD17, ZZ2 |
|  | Rice, pkt white | 2010-23 | na | AD21, ZZ1 |
|  | Bread, unsliced white loaf / Bread | 2010-23 | na | AD24, ZZ3 |
| **Other less healthy foods** | Bongos | 2010-23 | na | AE47, ZZ38 |
|  | Raw sugar, loose | 2010-23 | na | AE32, ZZ31 |
|  | Condensed milk, sweetened, tin | 2010-23 | na | AC28, ZZ23 |
|  | Cake sponge roll, length or wt / Cake | 2010-23 | na | AD27, ZZ6 |
|  | Buns, sweet plain / Buns | 2010-23 | na | AD25, ZZ4 |

#### Table D: Identification of potential substitute foods in the Customs dataset

| **Food product** | **HS codes and descriptions** |
| --- | --- |
| Untaxed meat | 160210 Meat preparations: homogenised preparations of meat, meat offal or blood  160231 Meat preparations: of turkeys, prepared or preserved meat or meat offal (excluding livers and homogenised preparations)  160239 Meat preparations: of poultry (excluding turkeys), prepared or preserved meat or meat offal (excluding livers and homogenised preparations)  160241 Meat preparations: of swine, hams and cuts thereof, prepared or preserved (excluding homogenised preparations)  160242 Meat preparations: of swine, shoulders and cuts thereof, prepared or preserved (excluding homogenised preparations)  160249 Meat preparations: of swine, meat or meat offal (including mixtures), prepared or preserved, n.e.s. in heading no.  160290 Meat preparations: of meat, meat offal or the blood of any animal, n.e.s. in heading no. 1602  Or in description:  'chicken',  ‘beef',  'pork',  'lamb',  'mutton',  'meat',  'chiken',  'turkey',  'duck',  'poultry',  'fowl',  ' hens',  'drumstick',  'wings',  'chick thigh'  'breast',  'tegel ch',  'tegel w',  'chick b',  'chkn ',  'lamp',  'sheep',  'chix ', |
| Crisps and snacks | 19030000  19042000  19043000  19059090 (excluding items with text: ‘cracker’, ‘biscuit’ or ‘cookie’) |
| Sauces | 2103 (excluding items with text: ‘mayo’) |
| Sugars | 180610 Hot chocolate  170111 Sugars: cane sugar, raw, in solid form, not containing added flavouring or colouring matter  170112 Sugars: beet sugar, raw, in solid form, not containing added flavouring or colouring matter  170113  170114  170191 Sucrose: chemically pure, containing added flavouring or colouring matter, in solid form  170199 Sucrose: chemically pure, not containing added flavouring or colouring matter, in solid form  170210 Sugars: lactose, chemically pure, in solid form: lactose syrup, not containing added flavouring or colouring matter  170211  170219  170220 Sugars: maple sugar chemically pure, in solid form: maple syrup, not containing added flavouring or colouring matter  170230 Sugars: glucose and glucose syrup, not containing fructose or containing in the dry state less than 20% by weight of fructose, the syrup not containing added flavouring or colouring matter  170240 Sugars: glucose and glucose syrup, containing in the dry state at least 20% but less than 50% by weight of fructose, the syrup not containing added flavouring or colouring matter  170250 Sugars: fructose, chemically pure, in solid form  170260 Sugars: fructose (excluding chemically pure fructose), in solid form, containing in the dry state more than 50% by weight of fructose: fructose syrup, not containing added flavouring or colouring matter  170290 Sugars: n.e.s. in heading no. 1702, including invert sugar  170310 Sugars: molasses, from sugar cane, resulting from extraction or refining of sugar  170390 Sugars: molasses, from sugar beet, resulting from extraction or refining of sugar |
| Other processed foods | 190230 Food preparations: pasta (excluding stuffed), cooked or otherwise prepared  190410 Food preparations: obtained by the swelling or roasting of cereals or cereal products  190490 Food preparations: cereal or cereal products (excluding maize), in grain form, pre-cooked or otherwise prepared  190510 Food preparations: crispbread, whether or not containing cocoa  200520 Vegetable preparations: potatoes, prepared or preserved otherwise than by vinegar or acetic acid, not frozen  190120 Food preparations: mixes and doughs for the preparation of bread, pastry, cakes, biscuits and other bakers' wares  190190 Food preparations: of flour, meal, starch, malt extract or milk products, for uses n.e.s. in heading no. 1901  190520 Food preparations: gingerbread and the like, whether or not containing cocoa |
| Rice | 1006 |
| Crackers | ‘cracker’ |
| Wholegrain cereals | 100620  110411  110412  110419  110421  110422  110423  110429  110430 |

Note: Waivered foods like fish, fruit and vegetables were not included.

#### Consideration of controlled analyses

Controlled analyses using geographical controls were not possible due to data availability. Also, rice imports were affected by major drop-offs at the time of the major tax interventions. For this reason, rice-controlled analyses were not reported (the majority would show increases in tax food imports relatively to the larger decline in rice imports).

### Further results

#### Figure A: Tax impacts on taxed indicator food prices and import volumes in Tonga

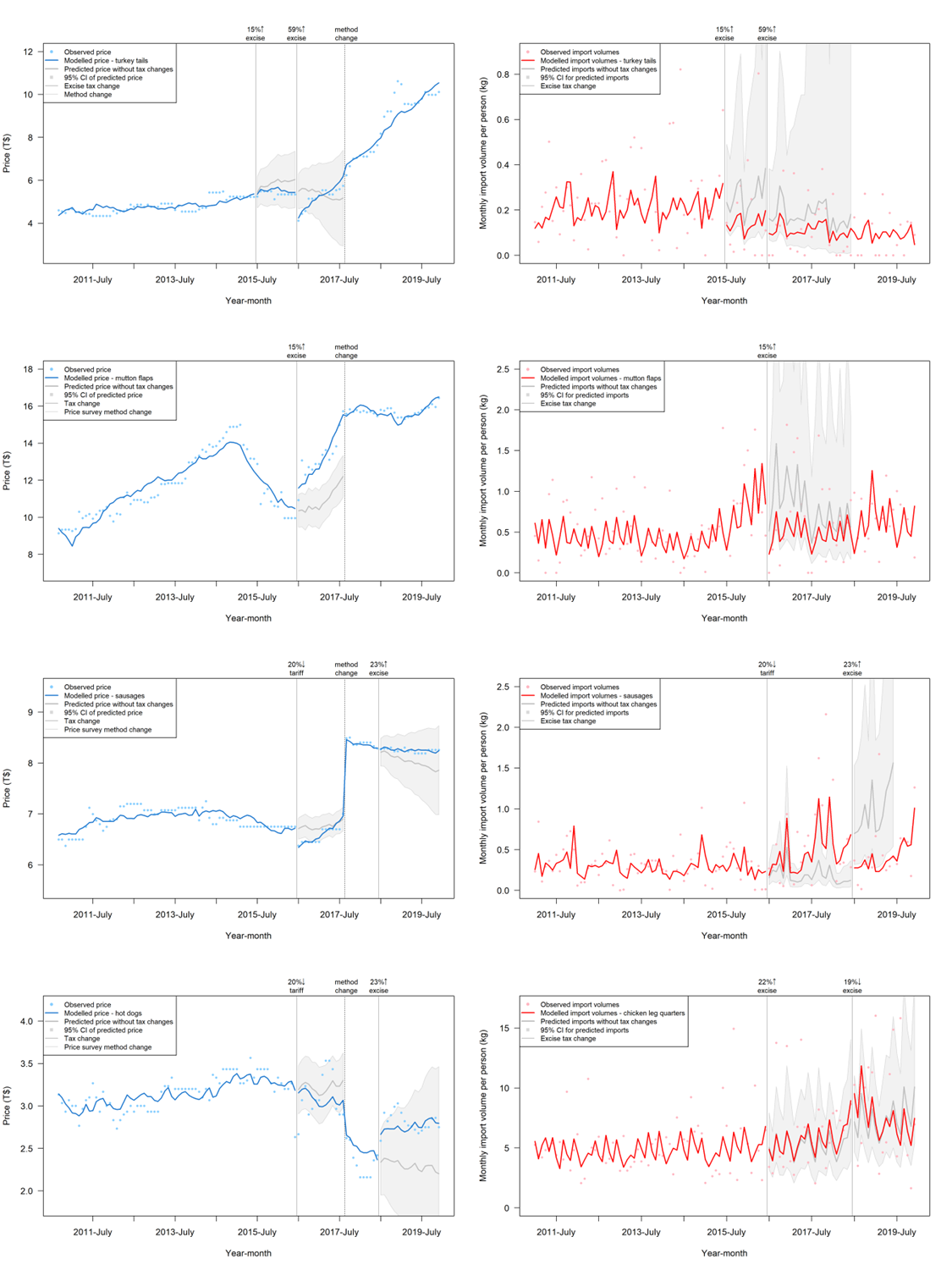

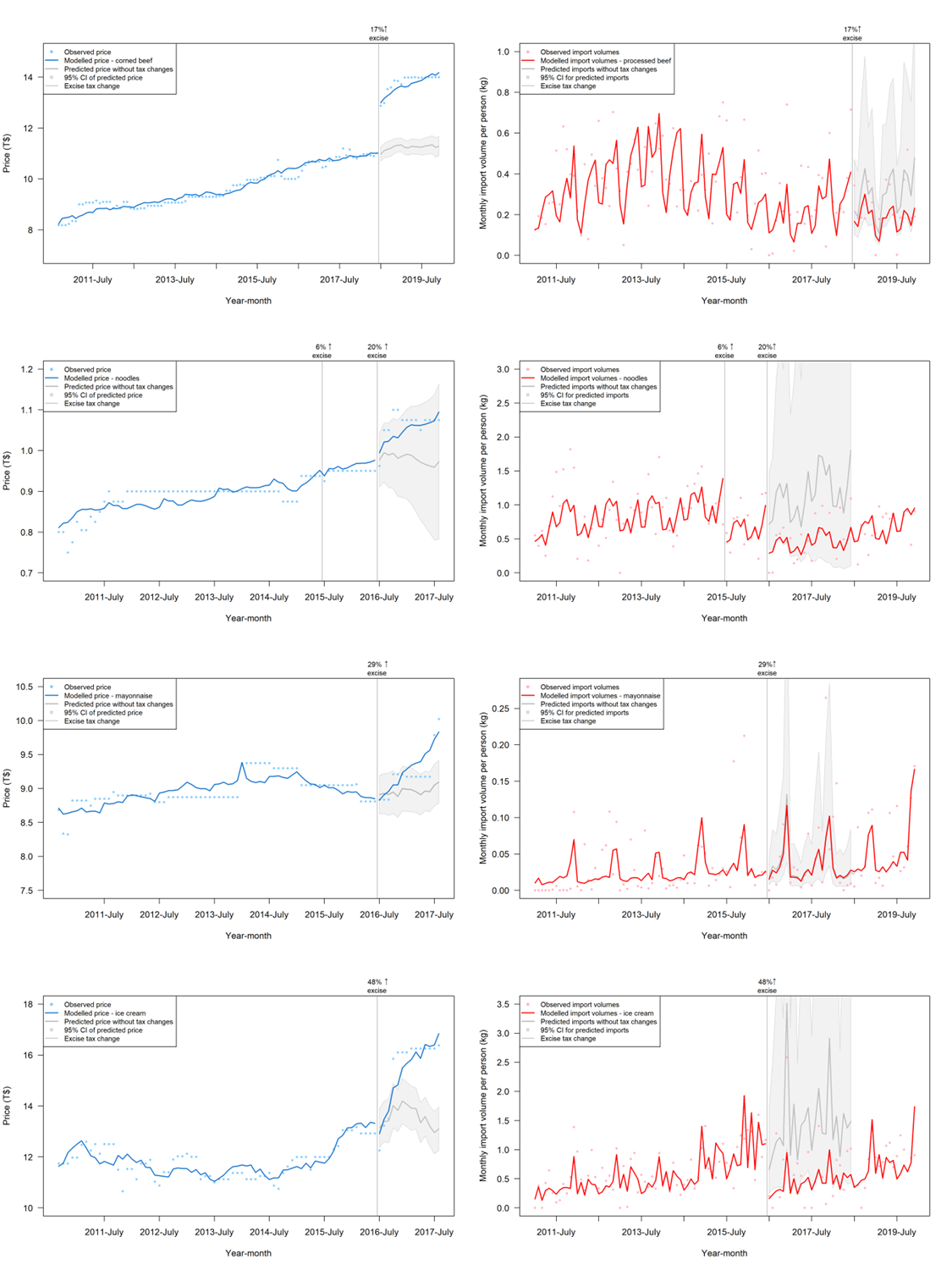

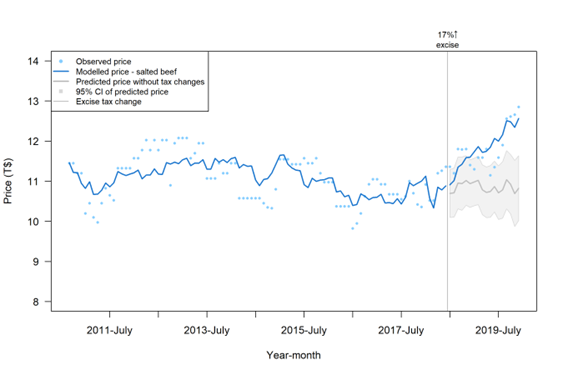

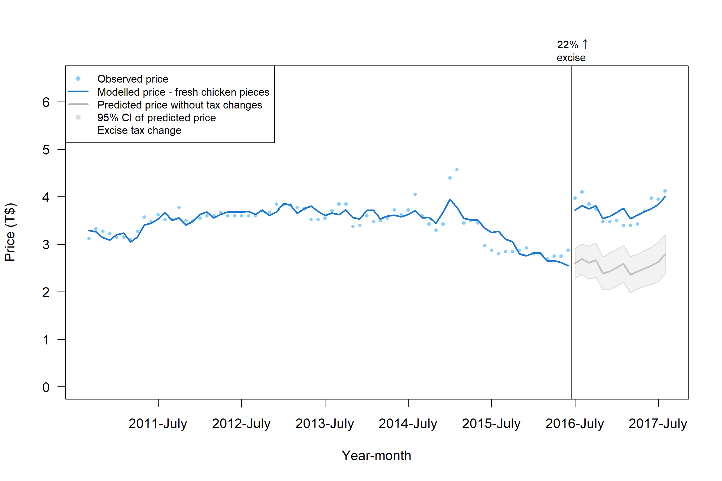

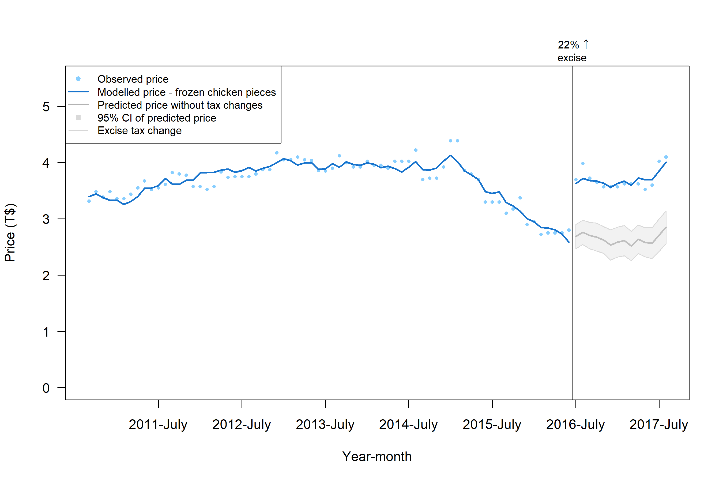

Notes: interrupted time series analysis adjusted for GDP, $T:$US exchange rate, international visitor numbers, month, oil prices (proxy of transport costs), and where relevant international prices of relevant food products such as chicken, beef, mutton and ice cream.

#### Figure B: Percentage change in imports in the two years post-tax increase, where two years follow-up was possible

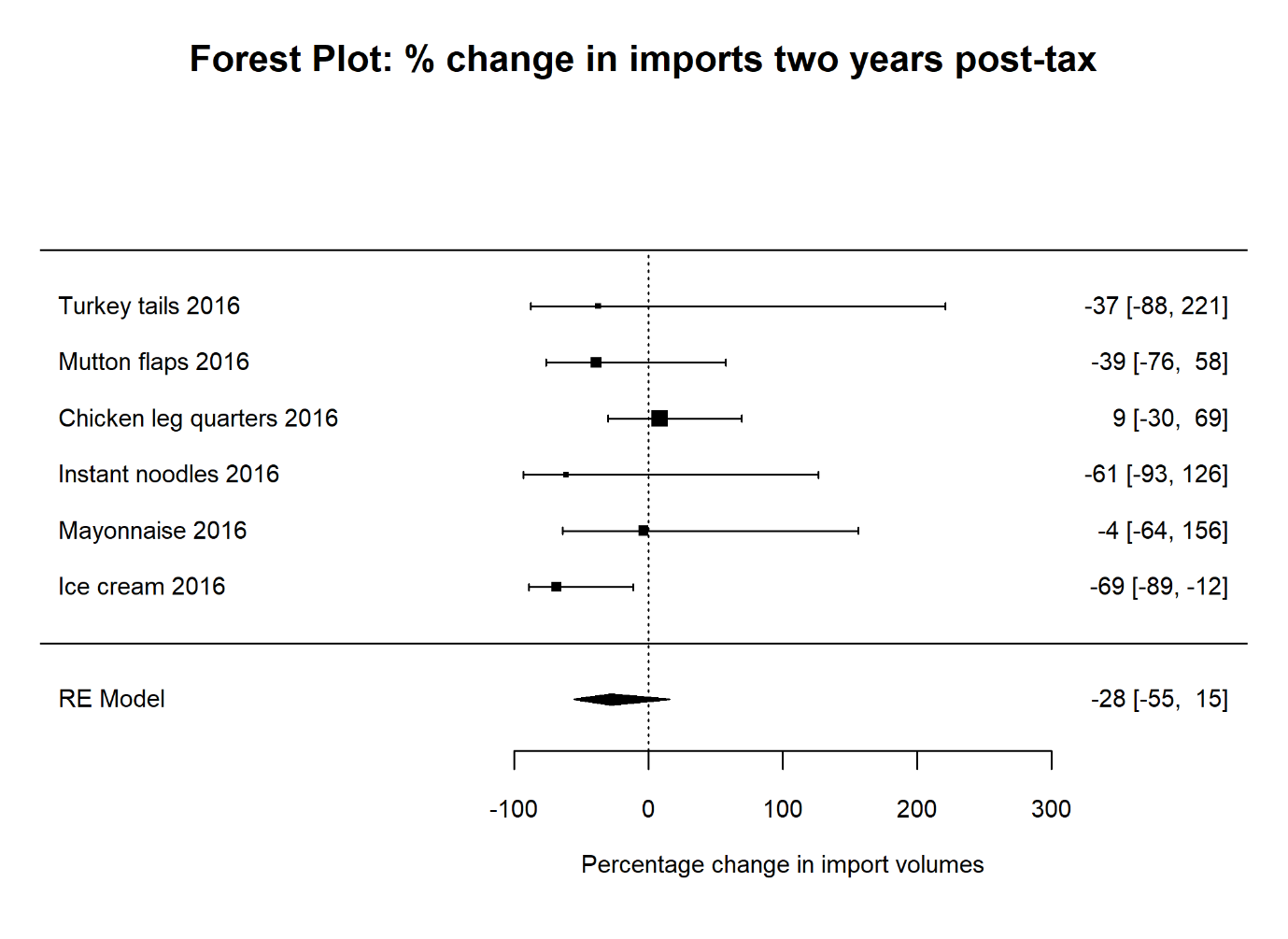

#### Table E: Total revenue changes as a proportion of import value (cost, insurance, freight inclusive) on imported foods targeted by taxes in Tonga

| **Food product** | **Year of tax** | **Tax increase** | **Revenue**  **pre-tax** | | **Revenue**  **post-tax** | | **Revenue**  **increase** | |
| --- | --- | --- | --- | --- | --- | --- | --- | --- |
|  | **change** | (AVE%) | (T$) | (AVE%) | (T$) | (AVE%) | (T$) | (% point △) |
| turkey tails | 2015 | 15 | 113,440 | 11 | 198,194 | 28 | 84,754 | 17 |
|  | 2016 | 59 | 198,194 | 28 | 355,196 | 128 | 157,002 | 99 |
| mutton flaps | 2016 | 15 | 941,449 | 15 | 2,063,658 | 32 | 1,122,209 | 17 |
| sausages | 2018 | 23 | 460,685 | 15 | 928,451^2^ | 41 | 467,766 | 26 |
| chicken leg quarters | 2016 | 22 | 1,734,110 | 14 | 5,247,845 | 33 | 3,513,735 | 19 |
| processed beef | 2018 | 17 | 824,918 | 15 | 1,238,608 | 31 | 413,690 | 16 |
| instant noodles^1^ | 2016 | 20 | 1,707,276 | 39 | 1,363,348 | 56 | -343,928 | 18 |
| mayonnaise | 2016 | 29 | 125,067 | 32 | 213,717 | 77 | 88,650 | 45 |
| ice cream | 2016 | 48 | 1,067,303 | 29 | 2,139,643 | 83 | 1,072,340 | 55 |
| **Total revenue** |  |  | 7,172,442 |  | 13,748,660 |  | 6,576,218 |  |

Note: 1. There was a decline in revenue from instant noodles at the same time there was a large reduction in imports (estimated at 60%). 2. An additional T$25,261 revenue was also collected one-year post-tax for locally manufactured sausages, not listed here. There was no revenue collected on locally manufactured ice cream and instant noodles in the first-year post excise tax. AVE%, ad valorem equivalent percentage, which is as a proportion of import value. Revenue is from tariff, excise and consumption taxes.

#### Table F: Pass-through estimates on imported foods targeted by taxes in Tonga. 2015-18

| **Food product** | **Year** | **Tax ↑** (T$/kg) | **Average price change**  (T$/kg) | **Pass-through**  (price change/tax increase) |
| --- | --- | --- | --- | --- |
| **Turkey tails** | 2015 | 0.48^4^ | -0.34 (-1.22 to 0.53) | -71% |
|  | 2016 | 1.50 | -0.29 (-1.40 to 0.82) | -19% |
| **Mutton flaps** | 2016 | 0.96^4^ | **2.12 (1.25 to 2.99)** | 221% |
| **Sausages** | 2018 | 1.00 | 0.16 (-0.22 to 0.55)  0.41 (-0.16 to 0.99)^1^ | 16%  41% |
| **Chicken leg quarters** | 2016 | 0.40 | **1.03 (0.80 to 1.27)^5^**  **1.16 (0.82 to 1.50)^6^** | 258%  290% |
| **Processed beef** | 2018 | 2.50 | **2.29 (2.01 to 2.56)**^2^  **0.71 (0.13 to 1.29)**^3^ | 92%  28% |
| **Instant noodles** | 2016 | 2.00 | 0.06 (-0.03 to 0.16) | 3% |
| **Mayonnaise** | 2016 | 2.00 | **0.26 (0.00 to 0.53)** | 13% |
| **Ice cream** | 2016 | 1.50 | **1.46 (0.71 to 2.20)** | 97% |
| **Average (mean)** | 2015-2018 | 1.37 | - | 68% |

Notes: 1. Price change for hot dogs; 2. corned beef; 3. salted beef; 4. These were increases in import tariffs, not excise taxes like the remainder. Tariff increases were converted into T$/kg using import unit value for the same year. 5. Price for frozen chicken pieces. 6. Price for fresh chicken pieces.

AVE is the ad valorem size of the tax change as a percentage of import value. GLS segmented linear model adjusted for GDP, month, international visitor numbers, exchange rates and oil prices and autocorrelation effects if relevant. Bolded results were statistically significant, p<0.05.

#### Figure C: Meta-analysis of excise tax effects on potential substitute food prices in the first-year post- 2016 and 2018 tax increases

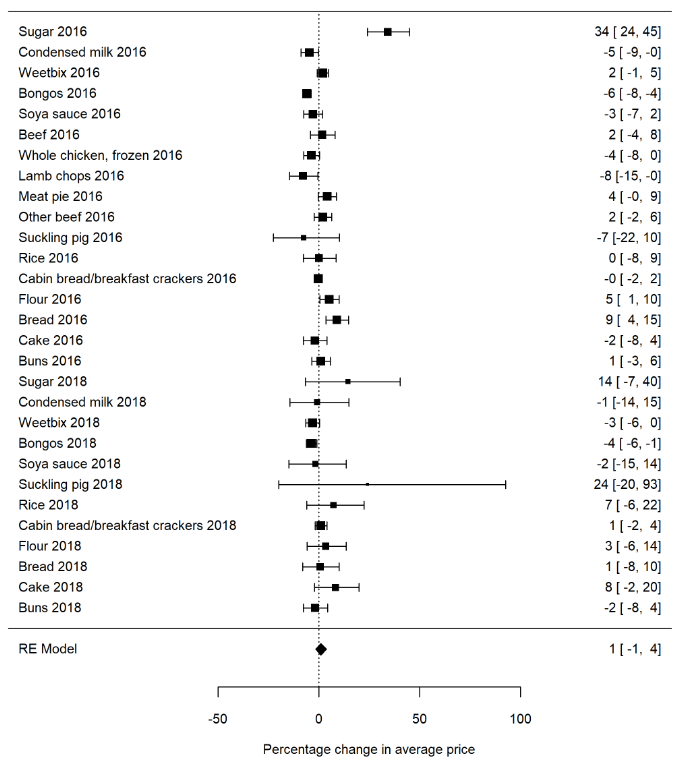

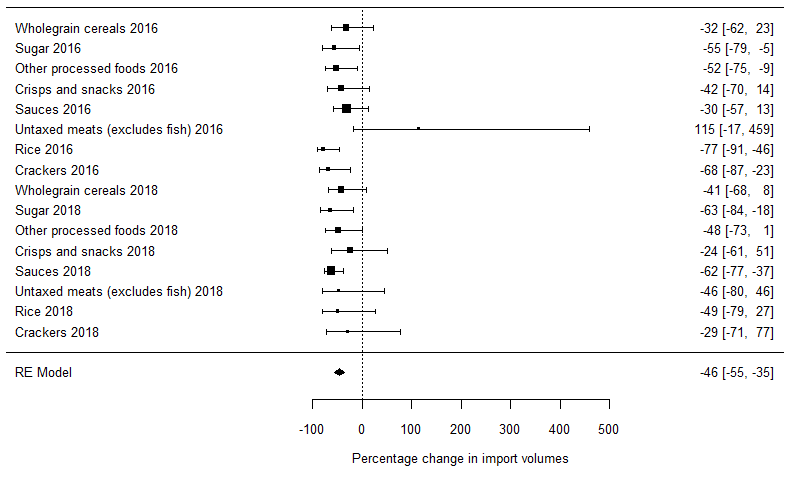

#### Figure D: Meta-analysis of excise tax effects on potential substitute food import volumes in the first-year post- 2016 and 2018 tax increases

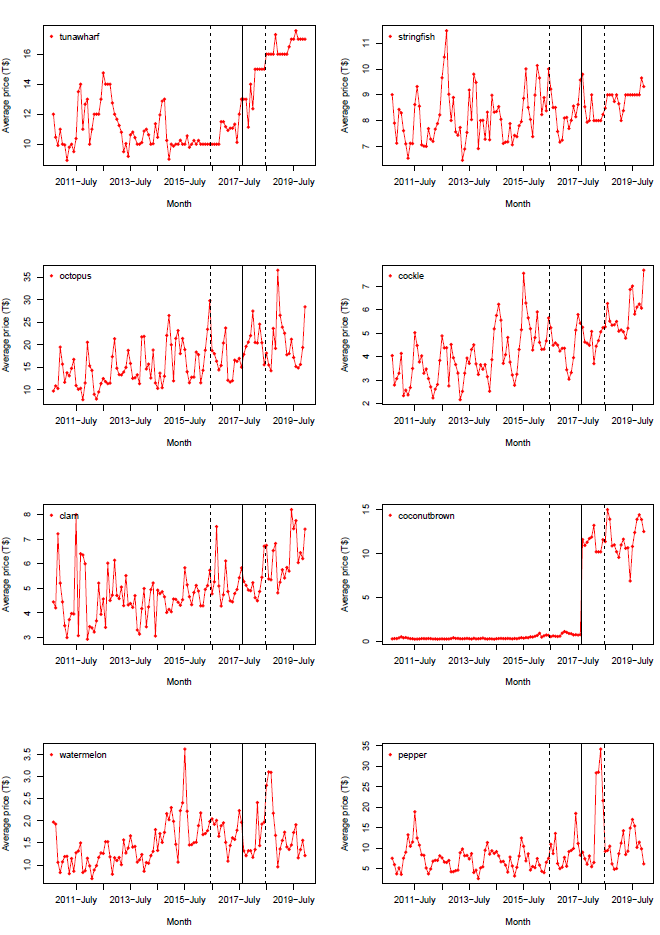

#### Figure E: Observed price trends for locally sourced foods over the time of the study period

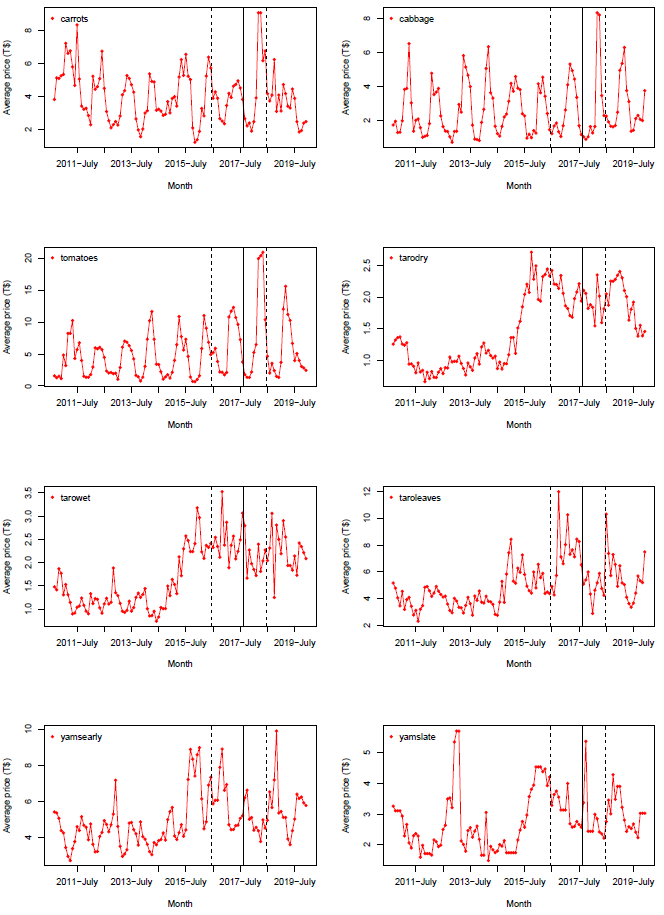

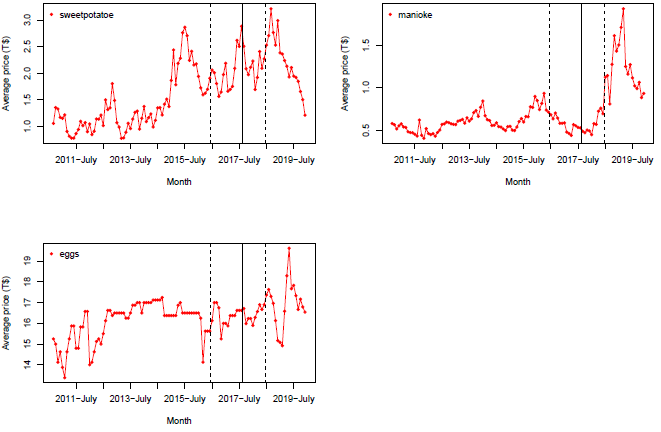
Notes: The dotted lines indicate the time of food tax increases. The straight line indicates a change in the method of data collection. All data sourced from retail surveys by Tonga Department of Statistics.

There appeared to be more locally produced foods with declines than increases in price in 2016, and more foods with increases than declines in 2018. Some locally produced staple foods appeared to have had stable prices over the whole study period eg, string fish, cabbage, carrots, and late yams. (Figure E)
